# Association of SARS-CoV-2 Infection and Cardiopulmonary Long COVID with Exercise Capacity and Chronotropic Incompetence among People with HIV

**DOI:** 10.1101/2023.05.01.23289358

**Authors:** Matthew S. Durstenfeld, Michael J. Peluso, Matthew A. Spinelli, Danny Li, Rebecca Hoh, Monica Gandhi, Timothy J. Henrich, Mandar A. Aras, Carlin S. Long, Steven G. Deeks, Priscilla Y. Hsue

## Abstract

**Background:** Long COVID has been associated with reduced exercise capacity, but whether SARS-CoV-2 infection or Long COVID is associated with reduced exercise capacity among people with HIV (PWH) has not been reported. We hypothesized that PWH with cardiopulmonary post-acute symptoms of COVID-19 (PASC) would have reduced exercise capacity due to chronotropic incompetence.

**Methods:** We conducted cross-sectional cardiopulmonary exercise testing within a COVID recovery cohort that included PWH. We evaluated associations of HIV, prior SARS-CoV-2 infection, and cardiopulmonary PASC with exercise capacity (peak oxygen consumption, VO_2_) and adjusted heart rate reserve (AHRR, chronotropic measure) with adjustment for age, sex, and body mass index.

**Results:** We included 83 participants (median age 54, 35% female). All 37 PWH were virally suppressed; 23 (62%) had prior SARS-CoV-2 infection, and 11 (30%) had PASC. Peak VO_2_ was reduced among PWH (80% predicted vs 99%; p=0.005), a difference of 5.5 ml/kg/min (95%CI 2.7-8.2, p<0.001). Chronotropic incompetence more prevalent among PWH (38% vs 11%; p=0.002), and AHRR was reduced among PWH (60% vs 83%, p<0.0001). Among PWH, exercise capacity did not vary by SARS-CoV-2 coinfection, but chronotropic incompetence was more common among PWH with PASC: 3/14 (21%) without SARS-CoV-2, 4/12 (25%) with SARS-CoV-2 without PASC, and 7/11 (64%) with PASC (p=0.04 PASC vs no PASC).

**Conclusions:** Exercise capacity and chronotropy are lower among PWH compared to SARS-CoV-2 infected individuals without HIV. Among PWH, SARS-CoV-2 infection and PASC were not strongly associated with reduced exercise capacity. Chronotropic incompetence may be a mechanism limiting exercise capacity among PWH.

## INTRODUCTION

Multiple studies have demonstrated that persons with HIV (PWH) have reduced exercise capacity compared to HIV-uninfected individuals.^1-7^ Mechanisms for reduced exercise capacity among PWH are unknown, but may include cardiac limitations given associations of HIV with atherosclerosis and heart failure, pulmonary limitations given associations of HIV with impaired diffusion capacity,^8,9^ sarcopenia,^10^ or other causes. A prior study that used combined cardiopulmonary exercise testing (CPET) and cardiac magnetic resonance imaging to evaluate exertional dyspnea in HIV did not identify differences in right or left ventricular contractile reserve or exercise-induced pulmonary hypertension, two previously hypothesized mechanisms.^11^

Our prior work and others suggest that having persistent cardiopulmonary symptoms after SARS-CoV-2 infection consistent with Long COVID is associated with reduced exercise capacity on CPET.^12,13^ Secondly, we have demonstrated that HIV infection is associated with Long COVID.^14,15^ In an exploratory analysis, we also found that HIV was associated with chronotropic incompetence.

Therefore, within a COVID recovery cohort that included PWH and included SARS-CoV-2 vaccinated and uninfected PWH, we sought to compare exercise capacity and chronotropy based on (1) HIV, (2) SARS-CoV-2 infection, and (3) among PWH, whether cardiopulmonary post-acute symptoms were associated with reduce exercise capacity or chronotropy.

## METHODS

### Study design

This was a cross-sectional substudy embedded within a San Francisco, California based COVID recovery cohort that included PWH including those with a history of SARS-CoV-2 infection with and without PASC and SARS-CoV02 vaccinated individuals without history of SARS-CoV-2 infection (NCT04362150).^16^

### Participants

We included adult participants over age 18 with diagnosed HIV in our cohort who had previously participated in an echocardiogram study visit and were able to participate in cycle ergometry and compared then to individuals without HIV who had already performed CPET in our cohort using the same research protocol. We excluded those with known cardiac disease including history of myocardial infarction, heart failure, atrial fibrillation, pulmonary hypertension, congenital heart disease, valvular heart disease, and those with severe pulmonary disease including those requiring home oxygen or with prior lung surgery. Finally, we excluded those with orthopedic, musculoskeletal, or neurologic issues that precluded participation in cycle ergometry.

### Exposures

The two primary exposures we studied were HIV infection and among those with HIV, SARS-CoV-2 coinfection stratified by presence of symptoms consistent with Long COVID at the time of CPET. Participants were classified as having never had SARS-CoV-2 infection if they reported no history consistent with symptomatic SARS-CoV-2 infection and no history of a positive SARS-CoV-2 test (including home testing) at the time of CPET. Participants were classified as having cardiopulmonary phenotype Long COVID if they had a confirmed SARS-CoV-2 infection and reported one or more new or worse symptoms including chest pain, shortness of breath, palpitations, fatigue, or reduced exercise capacity that persisted at least 90 days after onset of infection without an alternative diagnosis in accordance with the WHO consensus definition ^17^.

### Cardiopulmonary Exercise Testing

We performed cardiopulmonary exercise testing using a cycle ergometer (Lode Corival CPET) with continuous metabolic cart measurements of gas exchange (Medical Graphics Corporation Ultima CardiO_2_), 12 lead ECG monitoring, blood pressure and pulse oximetry measurement in accordance with guidelines.^18,19^ Work was increased at a set rate per minute (5-30 W/min rounded to nearest 5) to target a 10-minute test estimated from each participant’s measured maximum voluntary ventilation and self-reported habitual exercise. Participants were encouraged to maintain a cadence of 50-60 cycles per minute and exercise to their maximum ability unless stopped prematurely for safety. We classified the reasons for exercise limitations as we have previously reported.^13^

### Outcomes

The primary outcome was exercise capacity (peak VO_2_) on maximal cardiopulmonary exercise testing. Because of the demographic differences between those with and without HIV, for the primary comparison between people with and without HIV we used the percent of predicted exercise capacity achieved using the Wasserman equations for prediction ^20^. Secondary outcomes included classification of patterns among those with reduced exercise capacity less than 85% predicted, relative peak VO_2_ in ml/kg/min, absolute peak VO_2_ in L/min, heart rate response at peak exercise using the continuous adjusted heart rate reserve (AHRR) calculated as [(HR_peak_-HR_rest_)/(220-age-HR_rest_)] and chronotropic incompetence defined as adequate effort measured using a respiratory exchange ratio>1.05, peak VO_2_ <85% predicted, AHRR <80%, and no alternative explanation for exercise limitation ^21^.

### Correlative data

We used previously assessed HIV characteristics including duration of HIV infection, nadir CD4 count (self-reported and verified from medical records if possible), current CD4 count, CD8 count, and CD4/CD8 ratio. Additionally, most participants had hsCRP previously measured, which we included as exploratory measures.^22^

### Statistical Analysis

First we described participant demographics and medical history by HIV status using number and proportion for categorical variables and median and interquartile range for continuous variables. For unadjusted analyses, Fisher’s exact test was used for categorical variables and t-tests for normally-distributed continuous variables, and for correlation between continuous measures Pearson’s correlation coefficients and p-values were reported. We used linear regression models to estimate the mean differences in peak VO_2_ and AHRR between those with and without HIV and adjust for possible confounders including age, sex, and body mass index. To check whether our findings were robust to our decision to exclude medical history, sensitivity analyses were performed including medical history. Primary consideration was given to the effect estimates and confidence intervals, but p values <0.05 were considered statistically significant for the primary outcomes. Interaction terms were considered potentially meaningful if p<0.10. Analyses were performed using STATA 17.1.

### Approval

The University of California San Francisco Institutional Review Board approved this study (IRB 20-33000), and all participants provided written informed consent prior to participation. This study was reported in accordance with the STROBE guidelines for observational studies.

## RESULTS

### Participant Characteristics

Of 69 PWH in our cohort who completed an echocardiogram study visit, 6 had significant cardiopulmonary disease or used wheelchairs and were excluded; Of 63 eligible, 37 PWH completed CPET. Of those, 23 had a history of SARS-CoV-2 infection at a median 15 months prior (IQR 15-17) and 11 (48% of SARS-CoV-2 infected) reported symptoms of cardiopulmonary PASC. We compared these to 46 HIV-uninfected controls with history of SARS-CoV-2 infection who completed CPET at a median 18 months after infection (IQR 16-20) of whom 28 (61%) reported symptoms of cardiopulmonary PASC. The median age was 54 years old and 29 (35%) were female (Table 1). Among PWH, the median duration with diagnosed HIV was 21 years (IQR 15-28), all were virally suppressed on antiretroviral therapy at the time of CPET, median CD4 count was 608 (IQR 370-736) and median CD4/CD8 ratio 0.92 (IQR 0.56-1.27). 95% of participants with and without HIV received at least one SARS-CoV-2 vaccine prior to CPET, with 90% in both groups having been vaccinated greater than 3 months earlier.

**Table 1.** Demographics, Medical History, SARS-CoV-2 and HIV Characteristics

|  | People with HIV (N=37) | People without HIV (N=46) |
| --- | --- | --- |
| Age, median [IQR] | 57 (53, 62) | 49 (40, 57) |
| Female Sex | 5 (14%) | 24 (52%) |
| Hispanic//Latinx | 11 (32%) | 8 (18%) |
| Black/African American | 7 (21%) | 1 (2%) |
| Asian/Pacific Islander | 0 | 6 (13%) |
| White | 16 (47%) | 30 (67%) |
| Hypertension | 16 (43%) | 6 (13%) |
| Diabetes | 6 (17%) | 3 (7%) |
| Asthma/COPD* | 2 (11%) | 10 (23%) |
| Ever Smoker* | 7 (47%) | 9 (24%) |
| Body Mass Index, kg/m<sup>2</sup> | 28.7±5.2 | 30.1±7.8 |
| Hospitalized for COVID-19 | 1/23 (4%) | 7/46 (15%) |
| Vaccinated at time of CPET | 35 (95%) | 44 (96%) |
| SARS-CoV-2 Infected | 23/37 (62%) | 46/46 (100%) |
| Long COVID Symptoms at CPET | 11/23 (48%) | 28/46 (61%) |
| Months since SARS-CoV-2 infection | 16.0 (14.5, 17.2) | 18.0 (16.3, 20.0) |
| Years since HIV Diagnosis | 21 (IQR 15-28) |  |
| Nadir CD4 Count (self-reported) | 228 (50, 408) |  |
| Current CD4 count | 608 (IQR 270-736) |  |
| Current CD8 count | 707 (IQR 559, 904) |  |
| Current CD4/CD8 Ratio | 0.92 (IQR 0.56-1.27) |  |
| Current ART Regimen |  |  |
| INSTI-based | 29 (78%) |  |
| NRTI-based | 2 (5%) |  |
| NNRTI-based | 1 (3%) |  |
| PI-based | 2 (5%) |  |

Note current ART regimen was missing for 3 participants. Abbreviations: INSTI= Integrase
Strand Inhibitor-Based; NRTI=Nucleoside/Nucleotide Reverse Transcriptase Inhibitor-Based;
NNRTI=Non-Nucleoside Reverse transcriptase inhibitor; PI=protease inhibitor

**Table 2.**
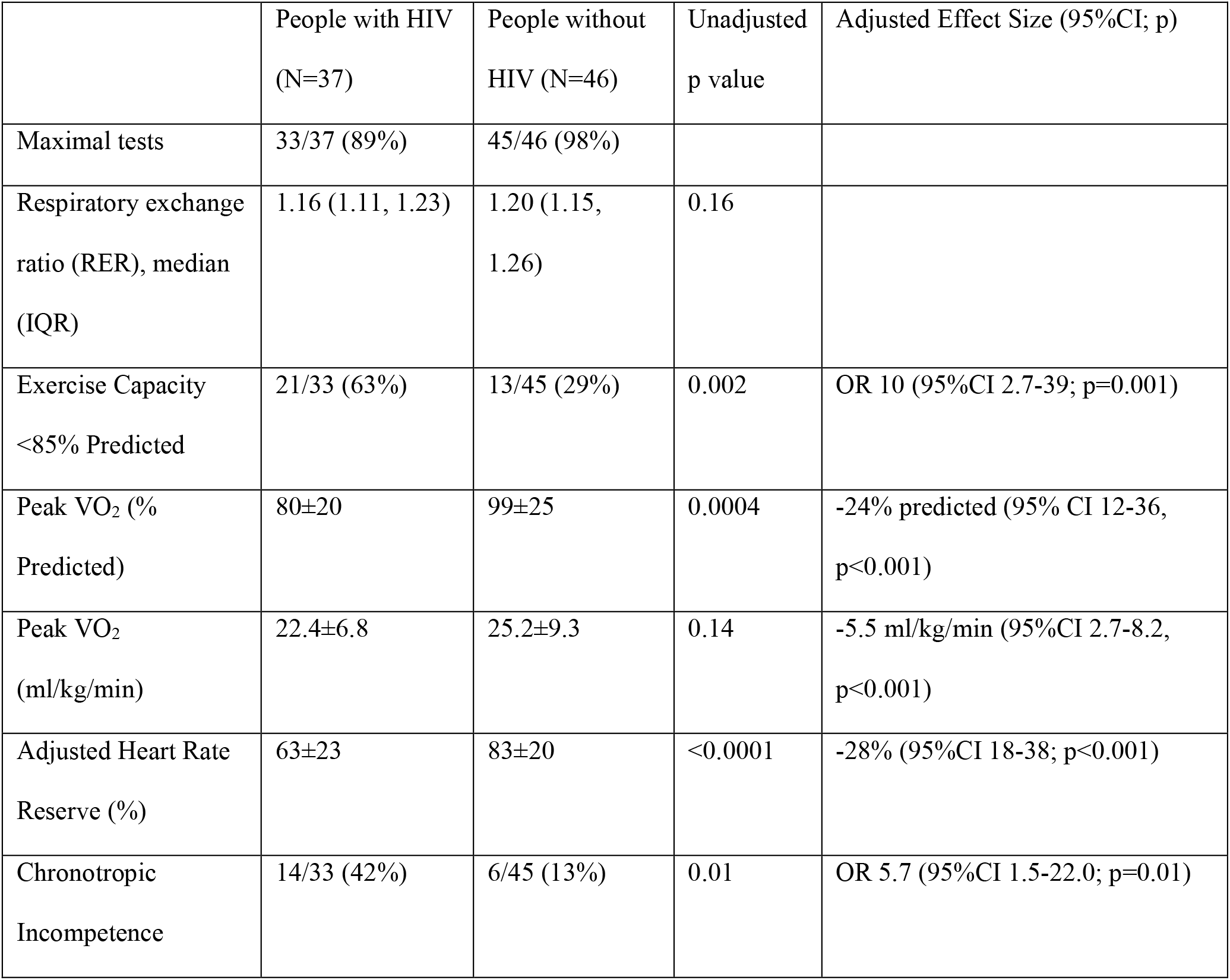
Key CPET Findings

|  | People with HIV<br>(N=37) | People without<br>HIV (N=46) | Unadjusted<br>p value | Adjusted Effect Size (95%CI; p) |
| --- | --- | --- | --- | --- |
| Maximal tests | 33/37 (89%) | 45/46 (98%) |  |  |
| Respiratory exchange<br>ratio (RER), median<br>(IQR) | 1.16 (1.11, 1.23) | 1.20 (1.15,<br>1.26) | 0.16 |  |
| Exercise Capacity<br><85% Predicted | 21/33 (63%) | 13/45 (29%) | 0.002 | OR 10 (95%CI 2.7-39; p=0.001) |
| Peak VO<sub>2</sub> (%<br>Predicted) | 80±20 | 99±25 | 0.0004 | -24% predicted (95% CI 12-36,<br>p<0.001) |
| Peak VO<sub>2</sub><br>(ml/kg/min) | 22.4±6.8 | 25.2±9.3 | 0.14 | -5.5 ml/kg/min (95%CI 2.7-8.2,<br>p<0.001) |
| Adjusted Heart Rate<br>Reserve (%) | 63±23 | 83±20 | <0.0001 | -28% (95%CI 18-38; p<0.001) |
| Chronotropic<br>Incompetence | 14/33 (42%) | 6/45 (13%) | 0.01 | OR 5.7 (95%CI 1.5-22.0; p=0.01) |

### Exercise Capacity among PWH compared to people without HIV

Exercise capacity was reduced among PWH compared to individuals without HIV with an achieved exercise capacity only 80% predicted among PWH vs 99% predicted among those without HIV (p=0.005), a difference in peak VO_2_ of 23.6 % predicted (95%CI 11.8 to 35.5; p<0.001) or 5.5 ml/kg/min (95%CI 2.7 to 8.2, p<0.001) after adjustment for age, sex, and body mass index. Results were similar when including history of diabetes and hypertension (19.5%, 95%CI 7.4-31.6; p=0.002) and diabetes, hypertension, and asthma/COPD (−15.9%, 1.7-30.1%, p=0.03). Among those with adequate effort (RER>1.05), exercise capacity was <85% predicted among 20/32 (64%) of PWH compared to 13/45 (29%) without HIV (unadjusted p=0.005; OR 10.9, 95%CI 2.7-44; p=0.001).

In stratified analysis by PASC vs no PASC, among those without PASC, 12/21 (57%) of PWH had reduced exercise capacity compared to only 2/18 (11%) without HIV (OR 52, 95%CI 1.4-1984; p=0.03). Among those with PASC, 8/11 (73%) of PWH compared to 11/27 (41%) without HIV (OR 6.7, 95%CI 1.05-43.1; p=0.04).

Among participants with HIV, 45% without SARS-CoV-2 coinfection had reduced exercise capacity compared to 73% with prior SARS-CoV-2 infection (unadjusted p=0.25; OR 1.7, 95%CI 0.30-9.9; p=0.54). Among those with HIV and SARS-CoV2 coinfection, the proportion with reduced exercise capacity did not vary by the presence of Long COVID symptoms (70% vs 73%; p=1.00; OR 3.5, 95%CI 0.78-15.5; p=0.10). Overall, the proportion with reduced exercise capacity among PWH (63%) was higher than the proportion with reduced exercise capacity among HIV-negative individuals with Long COVID (41%) and much higher than the HIV-negative SARS-CoV-2 recovered group (11%).

Compared to people with prior SARS-CoV-2 infection without HIV or Long COVID symptoms, PWH without SARS-CoV-2 coinfection achieved an exercise capacity 33% lower on the percent-predicted scale (95%CI -15 to -51; p=0.001), PWH with SARS-CoV-2 coinfection without Long COVID symptoms 26% lower among (95%CI -8 to -45; p=0.006) and PWH with Long COVID symptoms also 26% lower (95%CI -8 to -45; p=0.005). In other words, exercise capacity was reduced relative to predicted exercise capacity to a similar degree among PWH regardless of SARS-CoV-2 co-infection or Long COVID.

### Chronotropic Incompetence is Common in HIV and may be increased in Long COVID

Chronotropic incompetence, or the inability to augment heart rate appropriately during exercise without an alternative pattern for exercise limitation, was present in 38% of PWH vs 11% without HIV (p=0.002). AHRR (normal >80%) was lower among PWH vs individuals without HIV (60% vs 83%, p<0.0001). AHRR was 26% lower among PWH compared to people who were HIV-negative SARS-CoV-2 infected accounting for age, sex, and body mass index (95%CI 15.8 to 35.3; p<0.001).

Among PWH, heart rate response varied by SARS-CoV-2 recovery status: namely, 3/14 (21%) without SARS-CoV-2 infection had chronotropic incompetence, 4/12 (25%) with SARS-CoV-2 without Long COVID, and 7/11 (64%) with Long COVID (p=0.04).

Other patterns of reduced exercise capacity among PWH included cardiac limitations (i.e., ECG diagnostic for ischemia) in 3 participants, deconditioning/obesity in 2 participants, and ventilatory, pulmonary vascular, and hypertensive limitations in 1 each respectively.

### Correlates of Reduced Exercise Capacity and Chronotropic Incompetence

Higher BMI was inversely associated with exercise capacity even on the relative scale (ml/kg/min; Figure 2). There was a multiplicative interaction between BMI and HIV (p_interaction_=0.07) with a smaller difference of -0.50 ml/kg/min per 1 kg/m^2^ increase in BMI among those with HIV (95%CI -0.15 to -0.85; p=0.006) and -0.87 ml/kg/min per kg/m^2^ among those without HIV (95%CI -0.67 to -1.08; p<0.001). On the percent predicted scale, BMI was only associated with reduced exercise capacity among those without HIV (−1.1% predicted per kg/m^2^; 95%CI -0.26 to 2.0; p=0.01), but not among those with HIV (0.2; 95%CI -1.3 to 1.7; p=0.81).

**Figure 1.**
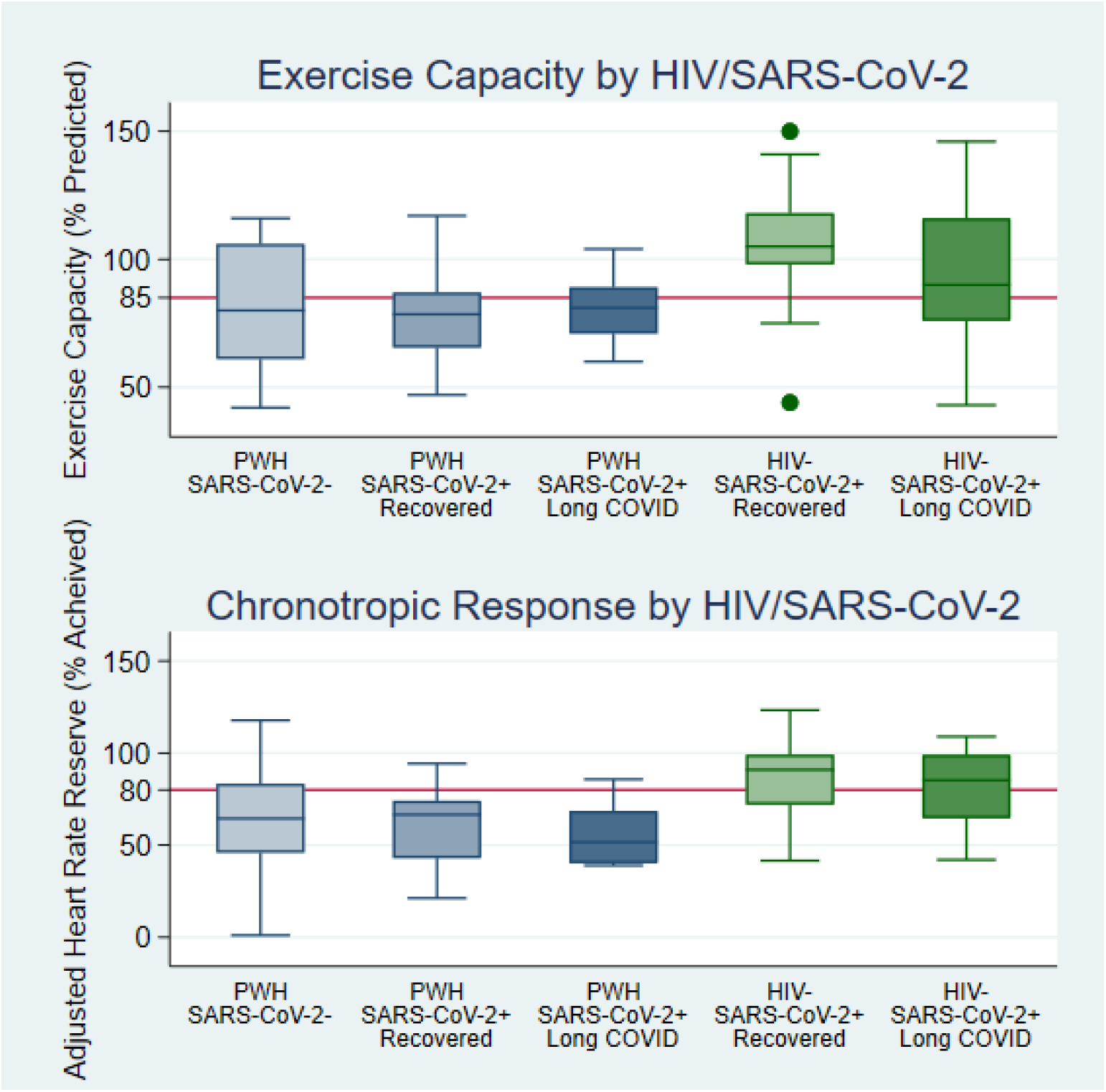
Exercise Capacity and Chronotropic Response by HIV and SARS-CoV-2 infection. Boxplots of exercise capacity on the percent predicted scale (top, red line indicates 85% which was our threshold for classifying as reduced) and adjusted heart rate reserve (bottom, normal >80%) are plotted by HIV and SARS-CoV-2 infection status. Exercise capacity is lower among PWH compared to individuals without HIV regardless of SARS-CoV-2 infection status. Adjusted heart rate reserve was lower among PWH, especially among those with symptoms consistent with Long COVID.

**Figure 2.**
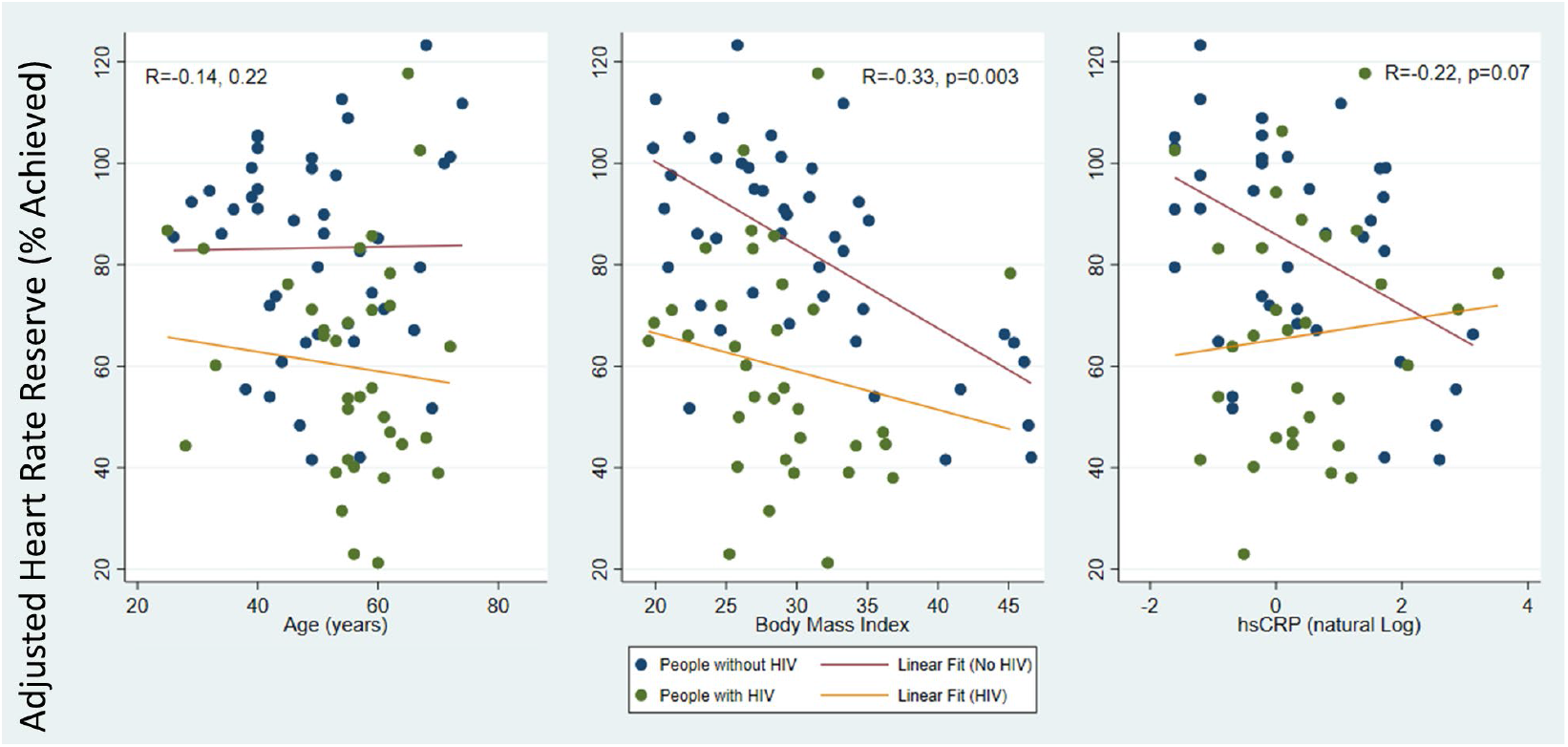
Chronotropy by Age, Body Mass Index, and hsCRP among those with and without HIV. Scatterplots with linear fit lines for adjusted heart rate reserve (AHRR, y-axis) by age (left), body mass index (center), and natural log transformed hsCRP (right) stratified by HIV status (green and orange for PWH, blue and red without HIV). Pearson correlation coefficients and p-values are for the unadjusted correlations for the total sample including those with and without HIV. The first panel demonstrates that AHRR is about 20% lower among PWH across the entire age spectrum compared to people without HIV. The second panel demonstrates that adjusted heart rate reserve decreases with increasing BMI with a stronger association among those without HIV, perhaps because individuals with low BMI start out with a lower AHRR compared to people without HIV. The third panel shows that although higher hsCRP is associated with lower AHRR among people without HIV, this is not evident among PWH.

Higher BMI was also associated with lower adjusted heart rate reserve (−1.4% per 1 kg/m^2^ increase in BMI; 95%CI -0.8 to 2.1; p<0.001) without significant interaction.

Similarly, higher hsCRP was associated with lower peak VO_2_ among people without HIV (−7.0% predicted per doubling, 95%CI -3.4 to -16.9; p=0.004) but not PWH (−1.2%, 95%CI -6.2 to 3.8; p=0.62; p_interaction_=0.06). Results were similar on the per kilogram scale (−1.3 ml/kg/min vs -0.7 ml/kg/min per doubling) although the interaction term was no longer statistically significant (p=0.38). We did not identify a multiplicative interaction between HIV and hsCRP on adjusted heart rate reserve, and hsCRP was not statistically significantly associated with adjusted heart rate reserve (2.3% per doubling; 95%CI -1.4 to 6.0; p=0.22).

In exploratory analyses among PWH, HIV disease specific characteristics including nadir CD4 count, current CD4 count, current CD8 count, and current CD4/CD8 ratio were not significantly associated with AHRR (Figure 3), although there was a trend toward a correlation between nadir CD4 count in the opposite of the predicted direction.

**Figure 3.**
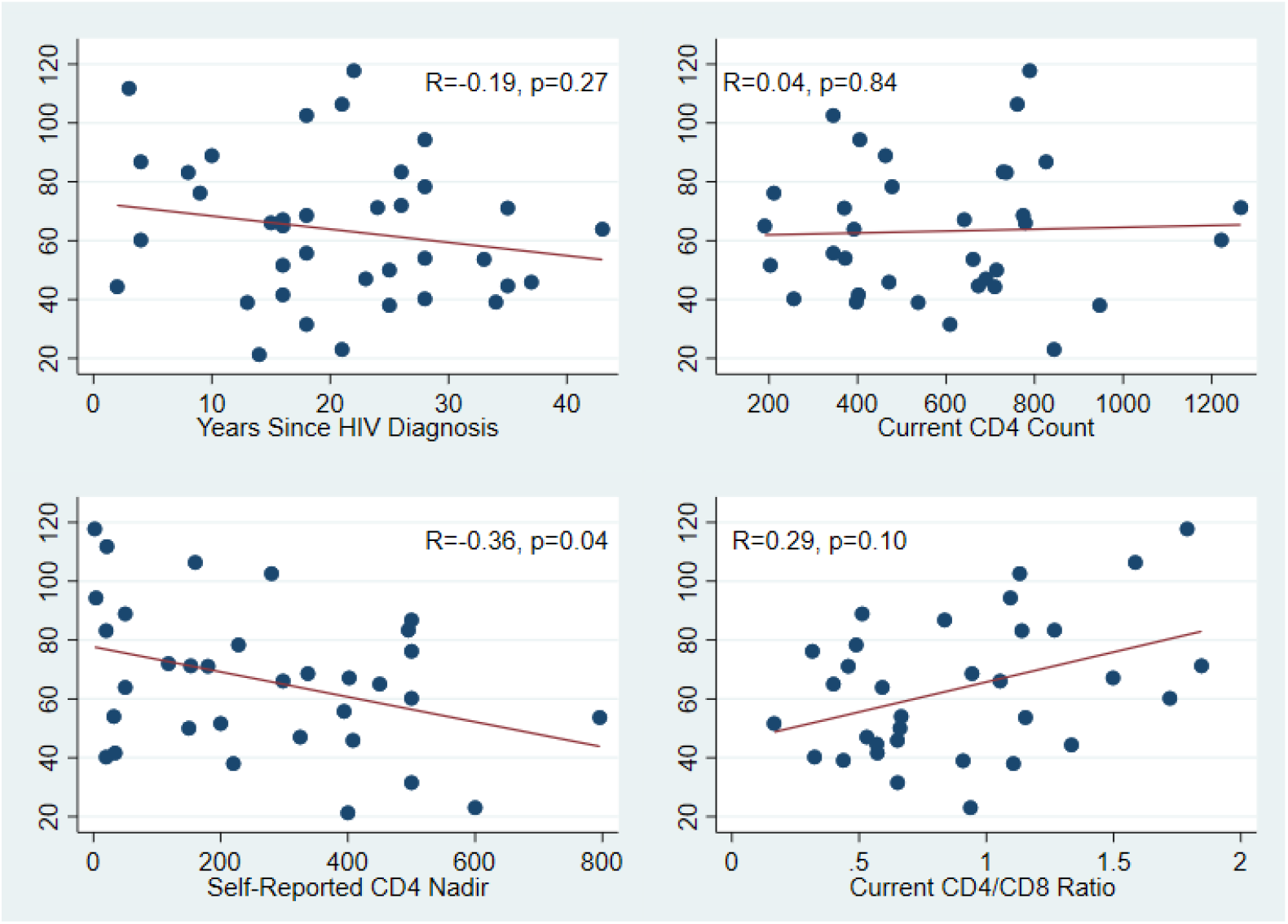
Chronotropy by HIV Characteristics. Adjusted heart rate reserve was not strongly correlated with years since HIV diagnosis, self-reported nadir CD4 count, current CD4 count, or current CD4/CD8 ratio, although there were trends of borderline statistical significance for self-reported CD4 nadir in the opposite direction we hypothesized and for current CD4/CD8 ratio. These exploratory analyses will need to be validated in larger studies.

## DISCUSSION

Reduced exercise capacity has been reported among people with HIV for 30 years ^1,23^, but few studies have explored the mechanisms of reduced exercise capacity. We found that exercise capacity was significantly reduced among PWH of similar magnitude to those with Long COVID, regardless of prior SARS-CoV-2 infection status, although our wide effect estimates lend significant uncertainty to this finding. Chronotropic incompetence was the most common pattern of exercise limitation we identified but did not explain all of the exercise limitations among PWH. Chronotropic incompetence was more common among those with HIV and SARS-CoV-2 coinfection experiencing cardiopulmonary symptoms compared to HIV-uninfected individuals with cardiopulmonary symptoms and PWH without symptoms.

### Consistent Findings Compared with Other Studies of Exercise Capacity in HIV

Our finding of reduced exercise capacity among PWH, though potentially subject to confounding given differences in baseline characteristics, is similar to prior reports. Prior studies have consistently identified that PWH have reduced exercise capacity compared to their peers who do not have HIV ^2^, even among newly diagnosed individuals as well as children and adolescents ^24,25^. Yet only a few studies have leveraged cardiopulmonary exercise testing to identify reasons for exercise limitations or exertional dyspnea. One earlier study found differences in peak arteriovenous oxygen differences suggestive of peripheral oxygen extraction or utilization limits ^26^. Higher body mass index, lipodystrophy and sarcopenia may also contribute.^4,10,27x^

### Contribution of SARS-CoV-2 Infection & PASC

Our study makes several novel contributions. To our knowledge, the effect of SARS-CoV-2 infection and PASC on exercise capacity has not been well-characterized, and our findings regarding chronotropic incompetence are interesting. The proportion of PWH we identified with chronotropic incompetence is similar to one prior study, which used treadmill stress tests rather than cardiopulmonary exercise testing, and found that 35% of PWH without known cardiovascular disease had chronotropic incompetence.^28^ They did not identify HIV-specific risk factors for chronotropic incompetence including duration of HIV, nadir CD4 count, or exposure to protease inhibitors.

### Potential mechanisms of chronotropic incompetence in HIV

The mechanisms of chronotropic incompetence in HIV, and specifically why the prevalence of chronotropic incompetence (and reduced exercise capacity more broadly) is so high among PWH remains unknown. One possible mechanism is that chronic inflammation from immune activation related to the underlying HIV viral reservoir may cause chronic adrenergic overactivation. Interestingly, a hyperadrenergic state is associated with elevated inflammatory markers including IL-6 in HIV.^29^ This, in turn, may result in reduced beta-receptor responsiveness, a common feature of chronotropic incompetence among people without structural heart disease.^30^ With decreased responsiveness to adrenergic signals, the natural response may be to generate higher levels of catecholamines in response to stress, which may activate inflammatory and hypercoagulable pathways that may accelerate atherosclerosis and cause cardiovascular events.

### Clinical significance of chronotropic incompetence

In the general population without HIV, chronotropic incompetence is a mechanism of reduced cardiopulmonary fitness associated with a particularly adverse prognosis. Data from multiple cohorts have demonstrated that chronotropic incompetence on stress testing among individuals without known cardiovascular disease or cardiopulmonary symptoms is associated with subsequent incident cardiovascular events including myocardial infarction and stroke, cardiovascular mortality, and all-cause mortality.^31-35^ Whether chronotropic incompetence has a causal role or is simply a marker of either poor cardiorespiratory fitness or underlying subclinical cardiovascular disease is uncertain among people without cardiovascular disease. To our knowledge, there are no data to inform whether chronotropic incompetence is associated with a similarly poor prognosis among PWH.

## Limitations

There are several limitations of this study. First, this is an observational study. In San Francisco, the prevalence of HIV is much higher among men than women, and many of our active research participants are older White men, limiting the diversity of our sample and possibly external generalizability. There were notable differences in baseline characteristics between those with and without HIV, including factors that may be associated with chronotropic incompetence, which is why we used the percent predicted exercise capacity rather than the absolute peak VO_2_ or relative peak VO_2_ as our primary measures for comparison between those with and without HIV. Although we conducted sensitivity analyses, there are still important confounders we could not adjust for including smoking status (given few current smokers among those without HIV) and unmeasured confounders such as pre-COVID-19 fitness. In terms of measurement, we did not perform invasive CPET to assess for differences in arteriovenous oxygen delivery or utilization or measure cardiac output, exercise diastolic function, or pulmonary hypertension with exertion, but we excluded those with evident structural heart disease on transthoracic echocardiography. Secondly, we did not confirm SARS-CoV-2 uninfected participants were uninfected by nucleocapsid antibody testing, so it is possible that some had previously had asymptomatic SARS-CoV-2 infection.

## Conclusions

Exercise capacity is reduced among PWH, with no large differences by SARS-CoV-2 infection or PASC, although our small sample size limits our ability to draw definitive conclusions. In contrast, we found that chronotropic incompetence may be a common and under-recognized mechanism of exercise intolerance among PWH especially among PWH with cardiopulmonary PASC similar to our findings among people without HIV.

## Data Availability

All data produced in the present study are available upon reasonable request to the authors.

## Notes

**Disclosures:** PYH has received modest honoraria from Gilead and Merck and research grant from Novartis unrelated to the submitted work. MJP has served as a consultant for AstraZeneca and Gilead Sciences, outside the submitted work. All other authors have declared that no conflict of interest exists.

**Funding:** This study was funded by philanthropic gifts from Charles W. Swanson, a grant from the NIH/NLBI (K12 HL143961), internal funds from the Division of Cardiology at Zuckerberg San Francisco General, and by a grant from the National Institutes of Health/NIAID (R01 AI158013). This work was supported by a UCSF-Gladstone CFAR Mentored Scientist Award via NIH grant to the UCSF-Gladstone Center for AIDS Research (P30AI027763). TJH is supported by NIH/NIAID 3R01A1141003-03S1. This publication was supported by the National Center for Advancing Translational Sciences, National Institutes of Health, through UCSF-CTSI Grant Number UL1TR001872. Its contents are solely the responsibility of the authors and do not necessarily represent the official views of the NIH.

### Competing Interest Statement

PYH has received modest honoraria from Gilead and Merck and research grant from Novartis unrelated to the submitted work. MJP has served as a consultant for AstraZeneca and Gilead Sciences, outside the submitted work. All other authors have declared that no conflict of interest exists.

### Funding Statement

This study was funded by philanthropic gifts from Charles W. Swanson, a grant from the NIH/NLBI (K12 HL143961), internal funds from the Division of Cardiology at Zuckerberg San Francisco General, and by a grant from the National Institutes of Health/NIAID (R01 AI158013). This work was supported by a UCSF-Gladstone CFAR Mentored Scientist Award via NIH grant to the UCSF-Gladstone Center for AIDS Research (P30AI027763). TJH is supported by NIH/NIAID 3R01A1141003-03S1. This publication was supported by the National Center for Advancing Translational Sciences, National Institutes of Health, through UCSF-CTSI Grant Number UL1TR001872. Its contents are solely the responsibility of the authors and do not necessarily represent the official views of the NIH.

### Author Declarations

The University of California San Francisco Institutional Review Board granted ethical approval for this study (IRB 20-33000), and all participants provided written informed consent prior to participation.

## REFERENCES

1. Pothoff G, Wassermann K, Ostmann H. Impairment of exercise capacity in various groups of HIV-infected patients. Respiration 1994;61:80–5.

2. Oursler KK, Sorkin JD, Smith BA, Katzel LI. Reduced aerobic capacity and physical functioning in older HIV-infected men. AIDS Res Hum Retroviruses 2006;22:1113–21.

3. Duong M, Dumas JP, Buisson M, et al. Limitation of exercise capacity in nucleoside-treated HIV-infected patients with hyperlactataemia. HIV Med 2007;8:105–11.

4. Gomes Neto M, Conceição CS, Ogalha C, Brites C. Aerobic capacity and health-related quality of life in adults HIV-infected patients with and without lipodystrophy. Braz J Infect Dis 2016;20:76–80.

5. de Lima LRA, Silva DAS, da Silva KS, Pelegrini A, de Carlos Back I, Petroski EL. Aerobic Fitness and Moderate to Vigorous Physical Activity in Children and Adolescents Living with HIV. Pediatr Exerc Sci 2017;29:377–87.

6. Orton PM, Sokhela DG, Nokes KM, Perazzo JD, Webel AR. Factors related to functional exercise capacity amongst people with HIV in Durban, South Africa. Health SA 2021;26:1532.

7. Robertson TE, Nouraie M, Qin S, et al. HIV infection is an independent risk factor for decreased 6-minute walk test distance. PLoS One 2019;14:e0212975.

8. Crothers K, McGinnis K, Kleerup E, et al. HIV infection is associated with reduced pulmonary diffusing capacity. J Acquir Immune Defic Syndr 2013;64:271–8.

9. Wang RJ, Nouraie M, Kunisaki KM, et al. Lung Function in Women With and Without Human Immunodeficiency Virus. Clin Infect Dis 2023;76:e727–e35.

10. Erlandson KM, Allshouse AA, Jankowski CM, MaWhinney S, Kohrt WM, Campbell TB. Functional impairment is associated with low bone and muscle mass among persons aging with HIV infection. J Acquir Immune Defic Syndr 2013;63:209–15.

11. Patterson AJ, Sarode A, Al-Kindi S, et al. Evaluation of dyspnea of unknown etiology in HIV patients with cardiopulmonary exercise testing and cardiovascular magnetic resonance imaging. J Cardiovasc Magn Reson 2020;22:74.

12. Durstenfeld MS, Sun K, Tahir P, et al. Use of Cardiopulmonary Exercise Testing to Evaluate Long COVID-19 Symptoms in Adults: A Systematic Review and Meta-analysis. JAMA Netw Open 2022;5:e2236057.

13. Durstenfeld MS, Peluso MJ, Kaveti P, et al. Reduced exercise capacity, chronotropic incompetence, and early systemic inflammation in cardiopulmonary phenotype Long COVID. J Infect Dis 2023.

14. Peluso MJ, Spinelli MA, Deveau T-M, et al. Postacute sequelae and adaptive immune responses in people with HIV recovering from SARS-COV-2 infection. AIDS 2022;36:F7–F16.

15. Peluso MJ, Deveau TM, Munter SE, et al. Chronic viral coinfections differentially affect the likelihood of developing long COVID. J Clin Invest 2023;133.

16. Peluso MJ, Kelly JD, Lu S, et al. Persistence, Magnitude, and Patterns of Postacute Symptoms and Quality of Life Following Onset of SARS-CoV-2 Infection: Cohort Description and Approaches for Measurement. Open Forum Infect Dis 2022;9:ofab640.

17. Soriano JM J; Diaz, JV; Murthy, S;; Relan, P. A clinical case definition of post COVID-19 condition by a Delphi consensus. In: Organization WH, ed. 2021.1 ed: ; 2021.

18. American Thoracic S, American College of Chest P. ATS/ACCP Statement on cardiopulmonary exercise testing. Am J Respir Crit Care Med 2003;167:211–77.

19. Balady GJ, Arena R, Sietsema K, et al. Clinician’s Guide to cardiopulmonary exercise testing in adults: a scientific statement from the American Heart Association. Circulation 2010;122:191–225.

20. Wasserman K HJ, Sue DY, Stringer W, Whipp BJ. Principles of Exercise Testing and Interpretation. 4th ed. Philadelpha: Lippincott Williams and Wilkins; 2005.

21. Brubaker PH, Kitzman DW. Chronotropic incompetence: causes, consequences, and management. Circulation 2011;123:1010–20.

22. Durstenfeld MS, Peluso MJ, Kelly JD, et al. Role of antibodies, inflammatory markers, and echocardiographic findings in postacute cardiopulmonary symptoms after SARS-CoV-2 infection. JCI Insight 2022;7.

23. Rigsby LW, Dishman RK, Jackson AW, Maclean GS, Raven PB. Effects of exercise training on men seropositive for the human immunodeficiency virus-1. Med Sci Sports Exerc 1992;24:6–12.

24. Somarriba G, Lopez-Mitnik G, Ludwig DA, et al. Physical fitness in children infected with the human immunodeficiency virus: associations with highly active antiretroviral therapy. AIDS Res Hum Retroviruses 2013;29:112–20.

25. Cade WT, Peralta L, Keyser RE. Aerobic capacity in late adolescents infected with HIV and controls. Pediatr Rehabil 2002;5:161–9.

26. Cade WT, Fantry LE, Nabar SR, Keyser RE. Decreased peak arteriovenous oxygen difference during treadmill exercise testing in individuals infected with the human immunodeficiency virus. Arch Phys Med Rehabil 2003;84:1595–603.

27. Vancampfort D, Mugisha J, Rosenbaum S, et al. Cardiorespiratory fitness levels and moderators in people with HIV: A systematic review and meta-analysis. Prev Med 2016;93:106–14.

28. De Lorenzo A, Meirelles V, Vilela F, Souza FC. Use of the exercise treadmill test for the assessment of cardiac risk markers in adults infected with HIV. J Int Assoc Provid AIDS Care 2013;12:110–6.

29. Robinson-Papp J, Astha V, Nmashie A, et al. Sympathetic function and markers of inflammation in well-controlled HIV. Brain Behav Immun Health 2020;7:100112.

30. Kawasaki T, Kaimoto S, Sakatani T, et al. Chronotropic incompetence and autonomic dysfunction in patients without structural heart disease. Europace 2010;12:561–6.

31. Lauer MS, Francis GS, Okin PM, Pashkow FJ, Snader CE, Marwick TH. Impaired chronotropic response to exercise stress testing as a predictor of mortality. JAMA 1999;281:524–9.

32. Myers J, Prakash M, Froelicher V, Do D, Partington S, Atwood JE. Exercise Capacity and Mortality among Men Referred for Exercise Testing. New England Journal of Medicine 2002;346:793–801.

33. Gulati M, Shaw LJ, Thisted RA, Black HR, Bairey Merz CN, Arnsdorf MF. Heart rate response to exercise stress testing in asymptomatic women: the st. James women take heart project. Circulation 2010;122:130–7.

34. Kodama S, Saito K, Tanaka S, et al. Cardiorespiratory Fitness as a Quantitative Predictor of All-Cause Mortality and Cardiovascular Events in Healthy Men and Women: A Meta-analysis. JAMA 2009;301:2024–35.

35. Jae SY, Heffernan K, Kurl S, et al. Chronotropic Response to Exercise Testing and the Risk of Stroke. Am J Cardiol 2021;143:46–50.

